# Does Recording Hardware Matter for Clinical Speech Recognition? Evaluating ASR Performance Across Consumer Devices

**DOI:** 10.64898/2026.05.19.26353590

**Authors:** Brian D. Tran, Di Hu, Seungjun Kim, Yawen Guo, Ramya Mangu, Tera L Reynolds, Jennifer Elston Lafata, Ming Tai-Seale, Kai Zheng

**Affiliations:** University of California, Irvine, Irvine, CA, USA; University of Maryland, Baltimore County, Baltimore, MD, USA; University of North Carolina at Chapel Hill, Chapel Hill, NC, USA; University of California, San Diego, San Diego, CA, USA

## Abstract

Ambient clinical intelligence (ACI) systems use automatic speech recognition (ASR) to capture patient-provider conversations for downstream clinical documentation. However, many ASR evaluations are conducted under controlled conditions using specialized hardware. We evaluated how recording devices influence transcription performance of contemporary ASR engines applied to clinical dialogue. Thirty-five primary care encounters were re-enacted from transcribed conversations and recorded using five devices simultaneously: smartphone, laptop microphone, portable recorder, clip-on microphone, and a desktop microphone. Six ASR engines were evaluated using word error rate (WER), clinical concept extraction precision and recall, and sentence-level semantic similarity. Median WER ranged from 16.7% to 20.7% across engines. Engine choice produced larger variation in transcription performance than recording device, although device-related differences were statistically significant. Overall, contemporary ASR engines demonstrated relative robustness to consumer-grade recording hardware, suggesting that model selection may have greater impact on transcription performance than recording device configuration in real-world ACI deployments.

## Introduction

Clinician burnout has been increasingly recognized as a public health concern^1,2^ with documentation burden considered as a major contributing factor^3,4^. To mitigate this burden, Ambient Clinical Intelligence (ACI), often referred to as “ai scribes”, has emerged as a promising approach to partially automate the documentation process^5,6^. These tools capture patient-provider dialogue using automatic speech recognition (ASR), after which natural language processing (NLP) and, more recently, specialized large language models (LLMs) generate clinical notes for integration into Electronic Health Record (EHR) systems^7–9^.

The field has evolved rapidly in recent years, with major vendors and health systems deploying integrated clinical workflow assistant products that combine ambient listening, note generation, and task automation (e.g., Microsoft Dragon Ambient Experience (DAX) Copilot, Solventum/3M Fluency Align, Abridge, among others) ^10,11^. As adoption accelerates, increasing attention has been turned towards safety, reliability, and generalizability of these systems across diverse clinical settings. Independent evaluations remain early, and the evidence base is still maturing with respect to long-term outcomes and downstream effects on care quality and reimbursement.

While ACI systems are evaluated across those multiple dimensions, transcription accuracy remains a foundational technical component. Because ASR converts patient-provider conversations into text for downstream processing, errors at this stage may propagate through subsequent NLP and LLM-based documentation pipelines^12^. ASR performance has historically been evaluated by measuring transcription accuracy, evaluated by Word Error Rate (WER)^13^. Substantial progress has been reported over the past decade. Early benchmarks recorded WERs as low as 11.6% using a dual-encoder architecture along with a sophisticated wall-mounted 16-channel beamforming microphone arrays in acoustically treated rooms^14^. More recently, transformer-based end-to-end architectures have further improved performance, achieving real-time WERs below 8% under optimal conditions^15^.

Despite these advancements, a significant gap persists between laboratory performance and real-world conditions. Many ASR evaluation studies are based on recordings generated in controlled environments using high-quality, professional-grade recording sites (e.g., sound-dampened exam rooms) with optimal device placement to maximize signal quality. While such setups support useful benchmarking for algorithm development, they may not reflect recording conditions in routine clinical practice.

In real-world settings, ACI systems are often implemented using more accessible recording device configurations, such as smartphones, laptops, tablets, or other consumer-grade devices. This shift toward Bring Your Own Device (BYOD) models introduces several sources of acoustic variability, such as background noise, gain inconsistency, and poor directional sensitivity^16^. If reliable transcriptions depend on specialized recording hardware this may limit the scalability of ACI systems in an equitable manner across high-volume ambulatory settings, community practices, and resource-constrained clinical environments.

Emerging usage patterns suggest that clinicians often rely on smartphones for recording and transcription, even though phones may need to be recharged between visits or, at times, during encounters^17^. However, these approaches may introduce implementation challenges, including dependence on compatible devices^18^, internet connectivity, potential privacy concerns, and possible disruption to the clinical encounter when recordings are managed outside integrated infrastructure^18^. Despite these realities, relatively little empirical research has examined how recording devices influence transcription performance of modern ASR systems applied to patient-provider dialogue^16^.

In this paper, we report an empirical study to evaluate how recording hardware affects the performance of contemporary ASR engines for clinical conversations. Our research questions were: 1) How does the choice of recording device (e.g., smartphone, laptop, portable digital recorder, or clip-on microphone) impact the transcription quality of state-of-the-art medical speech models applied to patient-provider dialogue, and 2) Does device configuration meaningfully affect preservation of clinically meaningful terminology or semantic content in resultant transcripts? By systematically comparing multiple recording devices and ASR engines using controlled recordings of clinical dialogue, this study provides empirical evidence on the robustness of contemporary speech recognition systems to variations in recording hardware, and offers practical insights for real-world deployment of ambient documentation technologies.

## Methods

### Conversation Data and Recording Device Configuration

The evaluation dataset for this study consisted of anonymized transcripts from 35 primary care encounters, representing 5 providers. The dataset was originally collected in an NIH-funded study that curated professionally transcribed and annotated clinical conversations in the exam room in primary care settings. This dataset has been previously used in several studies to investigate topics such as the discussion topic detection^19^, clinician adherence to best practices^20^, shared decision making^21^.

To control for potential undesired effects of recording-related factors (e.g., variations in volume and background noise) and speaker-related factors (e.g., non-native English speakers, pronounced accents), we re-enacted the dialogues. Two American English-speaking graduate students, one female and one male, read the de-identified transcripts in a quiet, sound-studio-like setting. A re-enactment guide, recording protocol, and ground-truth transcript development guide were generated to standardize evaluation data.

To study whether the quality of the recording devices and their relative placement between the two speakers may have an impact on ASR results, we used several different devices when re-enacting the dialogues. These devices are a wide range of affordable, everyday recording devices, many of which are already commonly used in clinical environments: 1) smartphone, 2) built-in microphone in laptop, 3) portable voice recorder, 4) clip-on microphone (worn by the physician speaker), and 5) medium-end (i.e., price range between $100 and $200) professional-grade desktop microphone. Specific models are detailed in Table 1. All devices, other than the clip-on microphone, were placed on a table approximately 1.5 feet to each of the speakers, configured to capture audio at lossless quality (Figure 1).

**Table 1.**
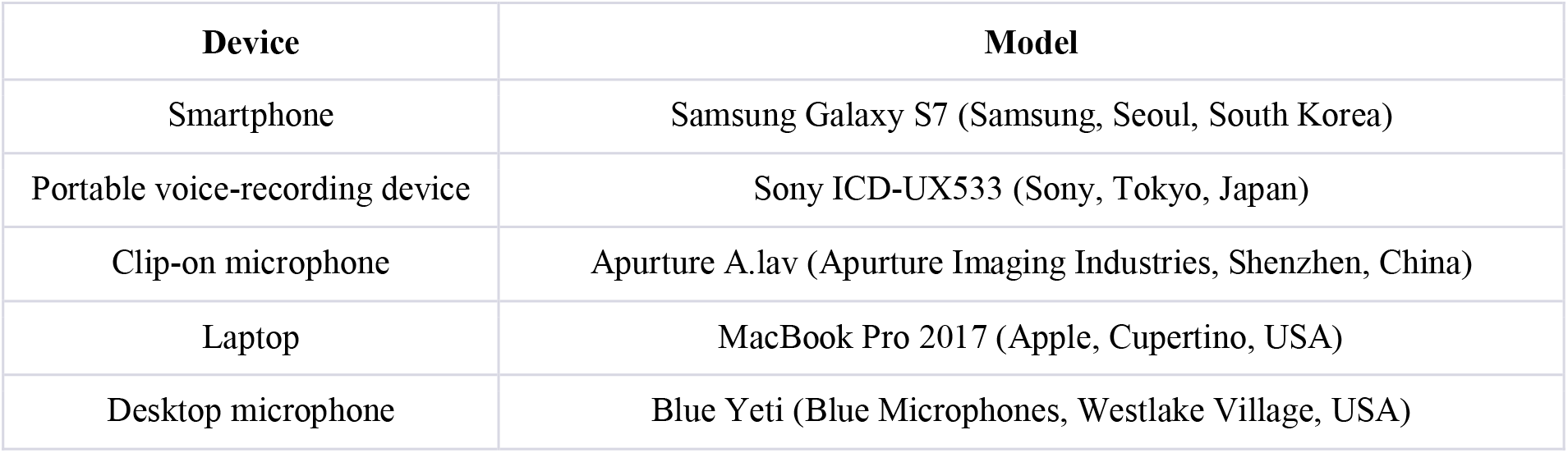
Recording devices used in the study.

**Figure 1.**
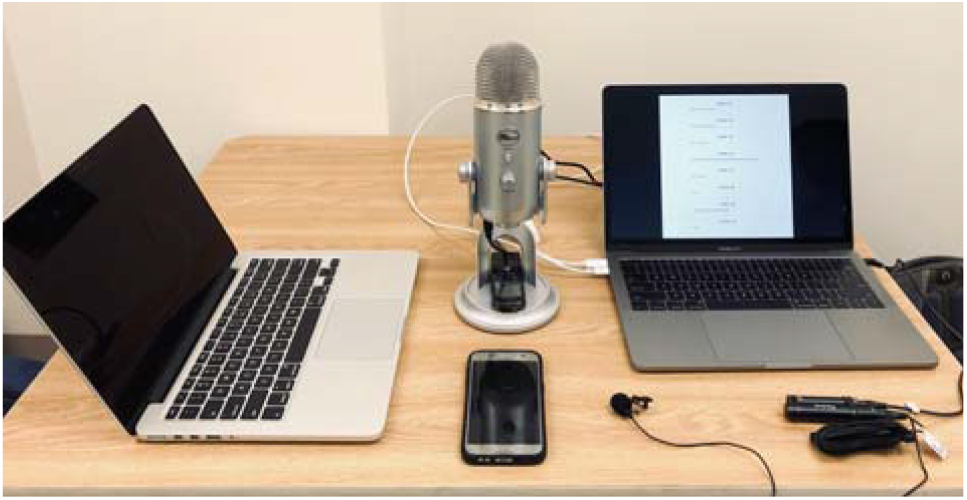
Arrangement of recording devices. Note: the computer to the right was used to show transcripts to actors. It was not used in recording. The Laptop recording device’s microphone area is encircled in red.

### Engine Selection and Evaluation Procedures

In this study, we selected six contemporary ASR engines, two of which were specifically designed for recognizing patient-provider conversations in the exam room. These included Google Speech-to-Text “medical_conversations” model and Amazon Web Services’ Transcribe Medical “primarycare” and “conversation” model, referred to as “Google Medical Conversation ASR” and “AWS Medical Conversation ASR” respectively. The other models included were Google “latest_long” model, AWS’s general transcribe’s model, Azure’s base model and “OpenAI Whisper model,” which we referred to as “Google General,” “AWS General,” “Azure General,” and “Azure Whisper” respectively. Transcription was completed in March 2026. While additional commercial digital scribe systems exist (e.g., AI DAX (Nuance, Burlington, USA; Abridge, Pittsburgh, USA), these platforms did not provide publicly accessible APIs for direct ASR evaluation and therefore were not included in the study.

We evaluated the transcription performance for this study using three contemporary metrics: WER, clinical concept precision and recall, and sentence-level semantic similarity. We used WER for straightforward comparison across existing ASR evaluation literature; this metric was defined as the number of word-level transcription errors (e.g., substitution, deletion, and insertion) over the total number of words in the reference transcript. We expressed the metric from 0 to 100%, with 0% (perfect transcription relative to reference), and 100% (completely different transcript). WER was calculated via the python package jiwer (3.0.4) with global alignment and package-associated transcript preprocessing^22^. Because WER does not necessarily capture whether clinically meaningful information is preserved, we supplemented the capture of biomedical concepts using named entity recognition. Clinical concepts were extracted via named-entity recognition using SciSpaCy (0.6.2) python package with the “en_core_sci_md” model^23^, and precision, recall, and F1-score were computed to assess omissions or incorrect insertions of medical terms such as clinical diseases. Finally, because word-level errors do not always alter broader sentence meaning (e.g., “I am sick” vs. “I’m sick”), we computed sentence level semantic similarity as a complementary measure of transcription fidelity. This metric captures preservation of general conversational meaning and is relevant as many ACI systems likely rely on downstream NLP or large language models to generate clinical documentation. Sentence-level embeddings were computed using the “sentence_transformers” (3.2.1) python package with the “all-MiniLM-L6-v2” model^24^.

### Statistical Analysis

ASR performance data such as WER are known to be skewed and to be influenced by a mixture of underlying error processes, resulting in non-normal distributions^25^. We therefore used non-parametric inferential statistical testing in our analysis, treating individual encounters as the unit of repeated measure. Friedman’s test for repeated measures was used to assess if performance metric distributions were significant across ASR engines and recording devices, with an α=0.05. Effect sizes were reported as Kendall’s W, which ranges from 0 to 1, with larger values indicating larger effect sizes. For interpretive purposes, values of approximately 0.1, 0.3, and 0.5 were considered small, moderate, and large effects, respectively. When omnibus testing was significant, post-hoc pair-wise comparisons were conducted via Wilcoxon-signed-rank tests with Holm-Bonferroni correction to account for multiple comparisons.

## Results

Overall, transcription accuracy varied across the six ASR engines tested (Table 2). Median WER ranged from 16.7% for AWS General to 20.7% for Google Medical. Significant differences in WER across engines were observed, with a very large effect size (χ^2^(5)=147.65, p=4.22e-30, W=0.84, n=35 encounters). Pairwise comparisons demonstrated statistically significant differences between all engine pairs (adjusted p < 0.05). These results indicate that engine choice, independent of recording hardware, can introduce meaningful variation in transcription accuracy.

**Table 2.**
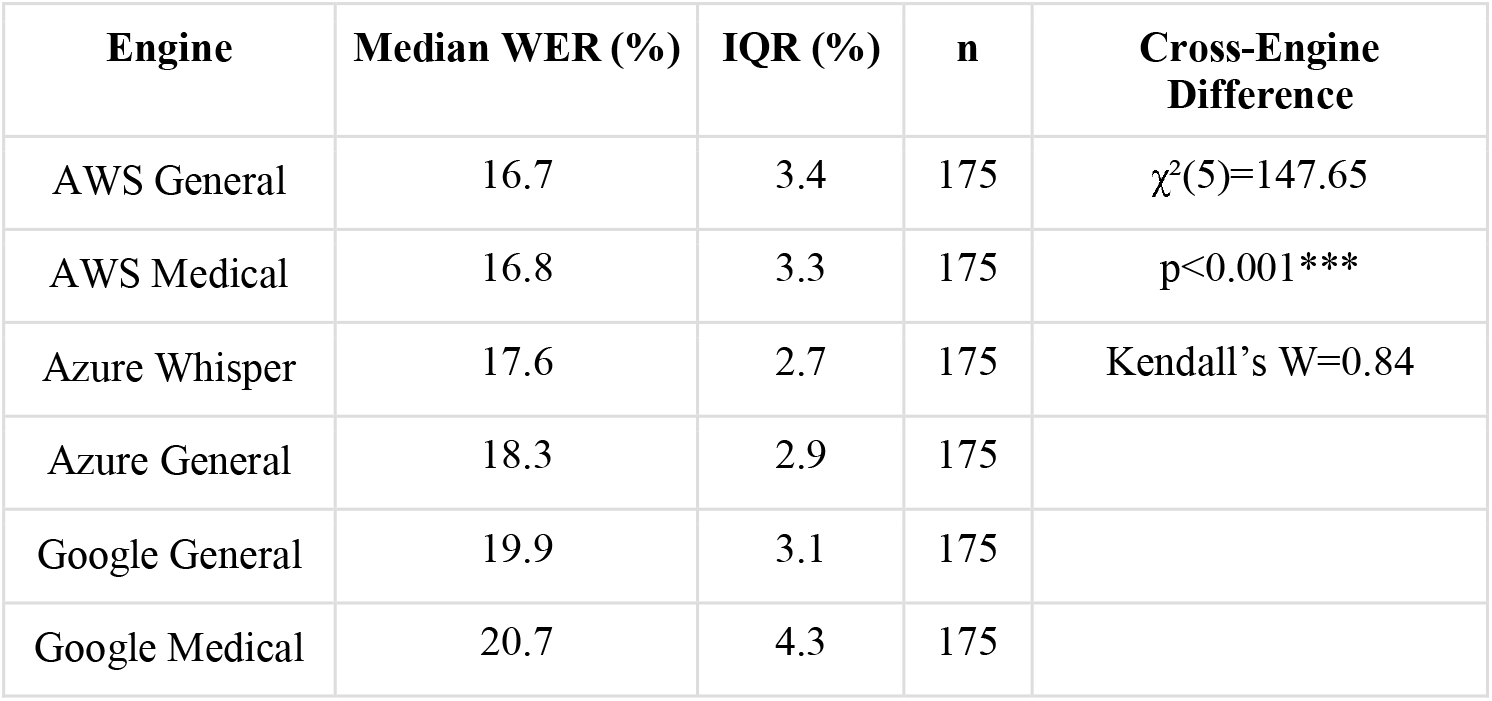
Baseline transcription performance by word error rate (WER) across 6 contemporary speech-recognition engines. Data is represented by 35 encounters and 5 recording devices, aggregated across recording devices recording each encounter simultaneously. Engines are ordered by increasing median WER (i.e., best performing to lowest performing). Cross engine differences were evaluated using Friedman test with effect size shown as Kendall’s W.

| Engine | Median WER (%) | IQR (%) | n | Cross-Engine Difference |
| --- | --- | --- | --- | --- |
| AWS General | 16.7 | 3.4 | 175 | $\chi^2(5)=147.65$ |
| AWS Medical | 16.8 | 3.3 | 175 | $p<0.001^{***}$ |
| Azure Whisper | 17.6 | 2.7 | 175 | Kendall’s $W=0.84$ |
| Azure General | 18.3 | 2.9 | 175 |  |
| Google General | 19.9 | 3.1 | 175 |  |
| Google Medical | 20.7 | 4.3 | 175 |  |

To evaluate the impact of recording modality on WER, WER distributions across devices were compared separately for each engine using a Friedman test (Table 3). Statistically significant device effects were detected across all engines, although effect sizes were smaller as compared to initial engine selection. For AWS models, Azure, and Whisper, device effects were small (W≈0.10-0.15). In contrast, both Google models showed larger device sensitivity (W≈0.46-0.48), indicating moderate variability in WER based on recording modality. Inspection of device-level distributions suggested that this effect was associated with the clip-on microphone configuration, which produced notably higher error rates relative to other recording devices for the Google models. These findings indicate the magnitude of device effect can vary across ASR engines.

**Table 3.** Effect of recording device on transcription on Word Error Rate (WER) for each ASR engine. WER distributions were compared using Friedman test, with effect size shown as Kendall’s W. WER values represent median WER (IQR) computed across 35 encounters with devices recording each encounter simultaneously. P-value <0.001 ***, <0.01 **, <0.05 *.

| Engine | n | Word Error Rate (%) |  |  |  |  | Cross-Device Difference |  |  |
| --- | --- | --- | --- | --- | --- | --- | --- | --- | --- |
| | | Desktop Median (IQR) | Recorder Median (IQR) | Cellphone Median (IQR) | Laptop Median (IQR) | Clip-On Median (IQR) | $\chi^2$ (df=4) | p-value | Kendall's W |
| AWS General | 35 | 16.7 (3.3) | 16.7 (3.3) | 16.8 (3.4) | 16.7 (3.6) | 16.8 (3.7) | 21.51 | <0.001*** | 0.154 |
| AWS Medical | 35 | 16.8 (3.2) | 16.8 (3.1) | 16.9 (3.2) | 16.8 (3.5) | 16.9 (3.6) | 21.18 | <0.001*** | 0.151 |
| Azure Whisper | 35 | 17.6 (2.5) | 17.9 (2.7) | 17.7 (3.1) | 17.6 (2.8) | 17.9 (2.8) | 14.13 | 0.007** | 0.101 |
| Azure General | 35 | 18.4 (2.6) | 18.3 (2.5) | 18.3 (2.8) | 18.3 (2.8) | 18.5 (3.0) | 13.49 | 0.009** | 0.096 |
| Google General | 35 | 19.7 (2.6) | 19.4 (2.5) | 19.6 (2.1) | 19.7 (2.9) | 22.0 (4.3) | 64.05 | <0.001*** | 0.458 |
| Google Medical | 35 | 20.6 (2.9) | 19.8 (2.9) | 20.0 (3.1) | 20.0 (3.3) | 25.0 (4.6) | 67.20 | <0.001*** | 0.480 |

We further characterize the impact of recording modalities by ranking WER by device (Figure 2A), and computing the range of within-encounter WER across recording devices for each engine (Figure 2B), which we denote as “device robustness.” Engines with smaller ranges could be interpreted as exhibiting greater stability across hardware configurations. With our device selection and evaluation dataset, the professional quality desktop microphone consistently produced the lowest WER. For device robustness, both Google models demonstrated larger variability in WER across devices with median device-range values were approximately 4%-5%, whereas the ranges were <3% for AWS and Azure models.

**Figure 2.**
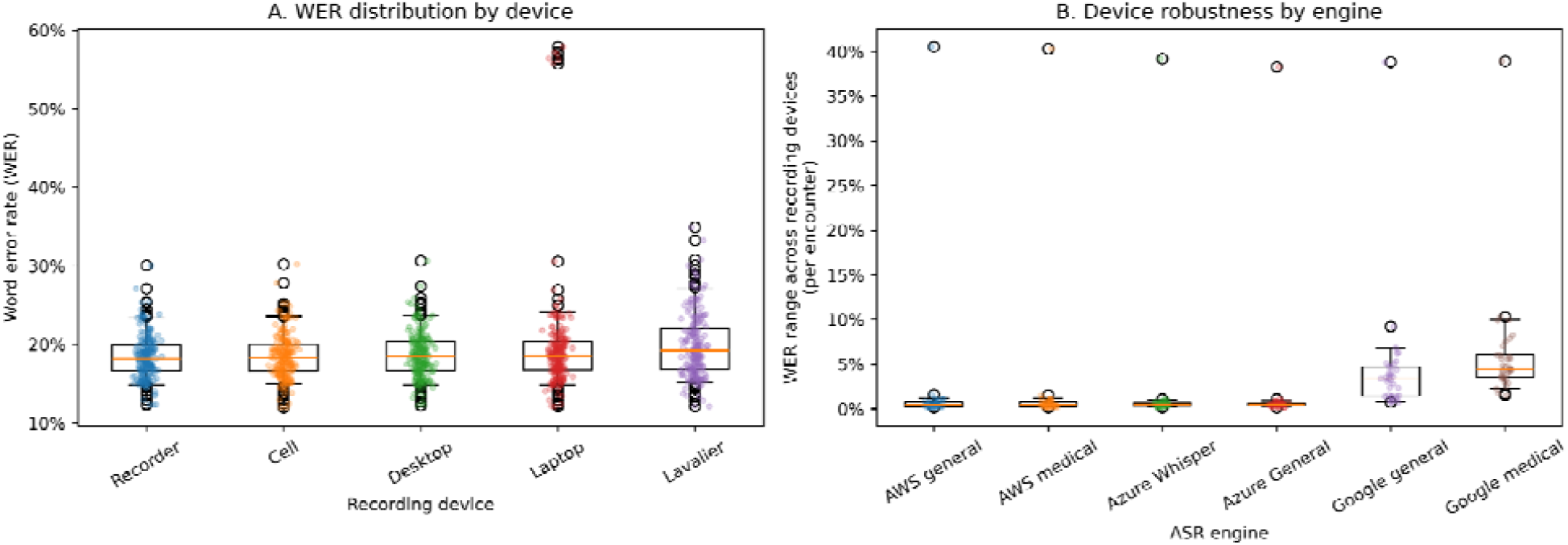
Impact of recording device on transcription accuracy across ASR engines: (A) Distribution of WER across 5 recording devices aggregated by 6 ASR engines. Each point represents the WER for a single encounter and engine combination. Boxplots show median and IQR. (B) Within-encounter range of WER across recording devices for each ASR engine. Larger ranges demonstrate greater sensitivity of an engine to variation in recording devices.

Inspection of WER distributions identified one encounter with unusually high WER across all ASR engines with the laptop microphone (WER≈56%). Recordings of the same encounter obtained from the other devices produced substantially lower error (WER<≈30%), suggesting a device-specific recording anomaly. To ensure that this outlier did not bias our findings, we performed a sensitivity analysis excluding this encounter and modality. The statistical significance of the device effect independently remained unchanged across engines.

Next, we examined whether recording devices affected the preservation of clinically meaningful terminology in the transcripts (Table 4). Device effects were evaluated separately for clinical term recall and precision using the Friedman test. Clinical term recall was significantly impacted by device choice for several of the evaluated engines. This was true particularly for the Google models (Google Medical: χ^2^(4)=55.86, p<0.001, W=0.40, Google General: χ^2^(4)=34.65, p<0.001, W=0.25). In contrast, AWS models showed relatively small recall effects (W≈0.07), and no significant device-related variation for Azure models. Precision of clinical terminology remained largely stable across most devices for most engines. The Google Medical models demonstrated a statistically significant, but small effect (χ^2^(4)=13.52, p<0.009, W=0.10). Other engines showed no significant differences. Overall, device-related variability was seen as reduced recall of clinical terminology rather than decreased precision.

**Table 4.**
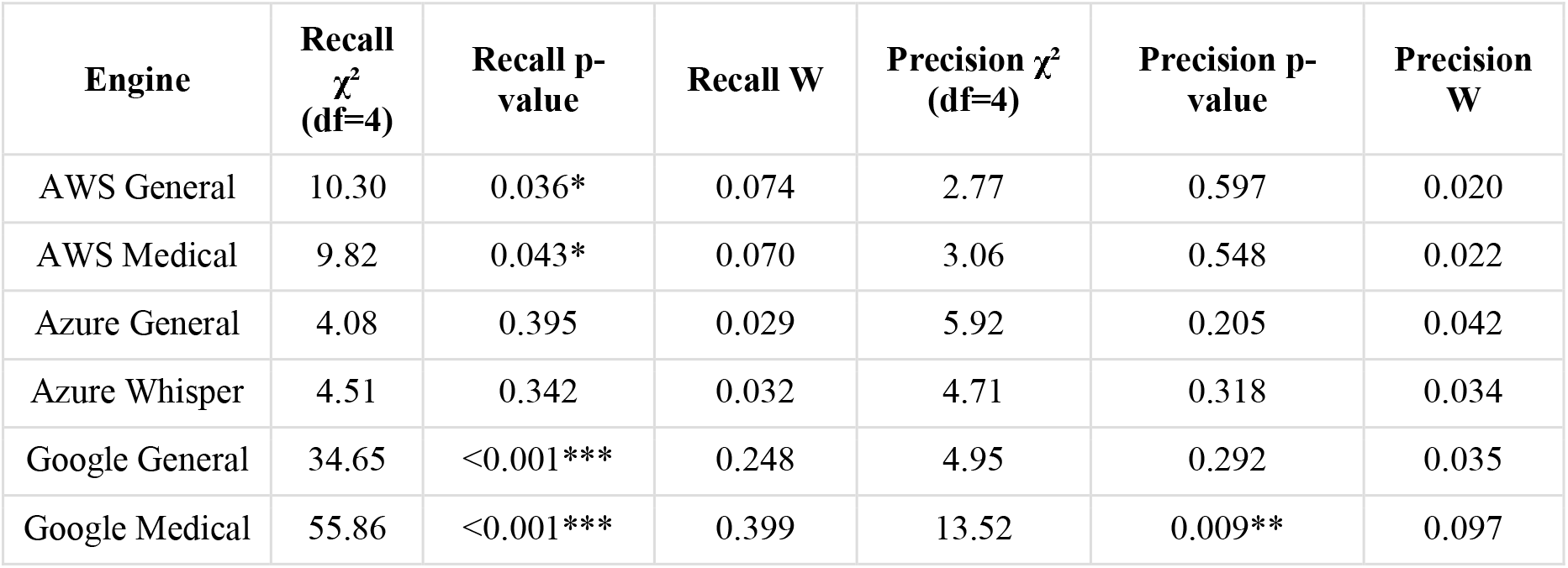
Effect of recording device on recall and precision of clinical terminology aggregated across devices for each ASR engine. Clinical concepts were extracted using SciSpaCy NER and compared with reference transcripts. A recording device effect was evaluated using Friedman tests for repeated measures, with effect sizes shown as Kendall’s W. p-value <0.001 ***, <0.01 **, <0.05 *.

**Table 5.** Comparison of sentence-level semantic similarity across recording devices for each ASR engine. Semantic similarity between ASR and reference transcripts was used to assess preservation of conversational meaning. A recording device effect was evaluated using Friedman tests for repeated measures, with effect sizes shown as Kendall’s W. p-value <0.001 ***, <0.01 **, <0.05 *.

| Engine | n | $\chi^2$ (df=4) | p-value | Kendall's W |
| --- | --- | --- | --- | --- |
| AWS General | 35 | 33.55 | <0.001*** | 0.240 |
| AWS Medical | 35 | 33.46 | <0.001*** | 0.239 |
| Azure General | 35 | 17.33 | 0.002** | 0.124 |
| Azure Whisper | 35 | 18.79 | <0.001*** | 0.134 |
| Google General | 35 | 32.07 | <0.001*** | 0.229 |
| Google Medical | 35 | 78.26 | <0.001*** | 0.559 |

To evaluate whether recording devices affected the preservation of overall conversational meaning, we also computed sentence-level semantic similarity between ASR and reference transcripts. Device-related variability was demonstrated for all engines, although the magnitude of this effect varied. Azure models showed relatively small device effects (W≈0.12-0.13), indicating relative preservation of semantic content across recording devices. AWS and Google General models demonstrated somewhat larger, but still small device effects (W≈0.22-0.24). In contrast, Google Medical showed notably greater variability in semantic similarity across devices (χ^2^(4)=78.26, p<0.001, W≈0.56), indicating that recording devices had a major effect on the preservation of transcript meaning. Overall, these findings indicate that devices introduce statistically detectable variation in the preservation of overall conversational meaning across ASR engines. However, the magnitude of this effect was generally small and mirrored the patterns seen in WER and clinical terminology recall.

## Discussion

In this study, we conducted a systematic evaluation of how recording hardware influences the performance of contemporary ASR engines applied to patient-provider dialogue. Across six engines and five recording devices, both engine selection and recording device affected transcription performance, although the magnitude of these effects varied. Variation across ASR engines was larger than the variation introduced by recording hardware, suggesting that transcription performance was driven more strongly by differences between ASR systems than by recording device alone. At the same time, recording devices introduced measurable differences in transcription quality across all engines. These effects were generally modest and not uniform across engines. Some engines demonstrated relatively strong robustness to device variation. These findings indicate that recording hardware configuration can have meaningful interactions with ASR models, producing variable transcription outcomes in otherwise identical conversations. Taken together, these results highlight that the performance of ASR systems used in ACI pipelines can be influenced by a combination of algorithmic and hardware-related factors. While selection of ASR models may yield the largest performance gains, recording hardware still plays a non-negligible role in real-world deployments.

Our observed WER values (≈16-21% across engines) fell within the range reported in prior evaluations of ASR applied to clinical conversations. Previous work has reported error rates ranging from 12% to 18% in large-scale conversational clinical benchmarks^14,26^. In smaller datasets, performance has varied depending on dataset characteristics, specialty, and recording conditions, such as with psychiatric consultations (≈25%) and accented or disordered speech (≈39-65%)^27–29^. Similar cross-engine differences have been reported, where notable variation was seen across vendors when ASR was evaluated on clinical dialogue tasks^30–32^. Together, these comparisons suggest that the observed performance is consistent with the current state of clinical conversational ASR.

The presence of device effects was consistent with prior work showing that microphone placement and environmental noise can materially affect ASR in clinical environments, including one study that evaluated a commercial clinical AI scribe^16^. While their work examined end-to-end performance of a single AI scribe system, our findings isolate the ASR stage and evaluates transcription fidelity across ASR engines.

Although device effects were smaller than engine effects, the magnitude of these effects varied across ASR systems. In this evaluation, the selected Google models demonstrated greater device-related variation in WER, clinical terminology recall, and semantic similarity relative to AWS and Azure models. Inspection of device-level results suggested this variability was driven in part by the clip-on microphone configuration for the Google Models. These differences suggest that ASR engines vary in their robustness to variability in recording hardware. One possible explanation is variation in the model training data; for example, models trained on more diverse audio sources, including consumer-grade microphones or mobile devices, may learn representations that generalize more effectively across different hardware configurations. While this study cannot assess model training, the observed differences highlight an important consideration for real-world ACI deployment. Systems intended for broad clinical adoption, where BYOD may be most practical, may benefit from models that are explicitly optimized for heterogeneous recording environments^14^.

Analysis of clinical concept extraction revealed a consistent pattern in how recording quality affected transcription errors. Device-related degradation of signal was associated with a reduced recall of clinical terminology, while precision remained stable across most engines and devices. Similar patterns have been observed in prior studies, where transcription errors more commonly result in omission of clinical terms rather than insertion of incorrect terminology^27,30,31^. The findings suggest that poorer recording conditions tended to result in omissions of medical terms rather than insertion of incorrect clinical concepts. This error pattern has important safety implications for downstream clinical documentation systems: omissions may reduce completeness or specificity of generated clinical notes, but insertion of incorrect clinical terms could introduce misleading or inaccurate information into the EHR^33^. Consistent with this interpretation, we also observed device-related effects on sentence-level semantic similarity across most engines, suggesting that the transcription errors did not substantially alter the overall conversational meaning. Because ASR output serves as input to downstream ACI pipelines, transcription errors may propagate into later stages of note generation^12^.

This study should be interpreted in the context of several limitations. First, the evaluation dataset consisted of re-enacted conversations derived from previously transcribed primary encounters. Although re-enactment allowed us to control for recording conditions, it may not fully replicate the natural complexity of patient-provider interactions. Real-world conversations may have speaker movement, varying accents and emotions, or overlapping speech which may influence ASR performance. Second, recordings were conducted in a quiet, sound-studio-like environment. While necessary to isolate the effects of recording hardware, it may underestimate the impact of environmental noise. Background conversations, medical equipment noise, and other environmental sounds may interact with recording devices in ways that were not captured in this evaluation. Third, the study evaluated a limited set of recording devices and placement configuration. Although we made an effort to select commonly-used consumer grade devices, additional configurations such as with wearable devices may benefit the generalizability of the findings. Fourth, the dataset included a limited number of encounters and speakers. A larger and more diverse sample of dialogues would similarly strengthen the generalizability of the findings. Finally, the evaluation focused on ASR transcription rather than the output of a complete digital scribe system. Many ACI solutions incorporate additional processing, such as error correction, speaker diarization, and LLM-based summarization, all of which may mitigate transcription errors before clinical notes are generated. As a result, the downstream impact on observed transcription differences on final documentation remains uncertain from this study alone.

Future research could extend this work in several directions. The use of real patient-provider encounters recorded in routine clinical environments will be useful to determine how recording hardware may interact with background noise, speaker movement, and diverse accents. Additionally, evaluating a broader range of devices would also help clarify how hardware influences transcription quality. A corollary to our findings is that future work should examine how ASR errors propagate through downstream ambient documentation pipelines, including systems which use LLMs. Understanding how transcription errors affect the final documentation accuracy, completeness, and clinical usability will be critical for evaluating the safety and effectiveness of ACI systems.

## Conclusion

This study evaluated how recording hardware influences the performance of contemporary ASR engines applied to patient-provider dialogue. Variation across ASR engines was substantially larger than variation introduced by recording devices, although device choice had a statistically detectable effect across WER, clinical concept recall, and sentence-level semantic similarity. Overall, recording hardware influenced transcription quality, but its impact was smaller than engine selection and varied markedly across ASR engines, indicating that hardware robustness can be an engine-specific property rather than a uniform characteristic of modern clinical ASR. Further evaluation in routine clinical settings is needed to understand how device variability affects downstream AI-assisted clinical documentation.

## Data Availability

All data produced in the present study are available upon reasonable request to the authors

## Funding and Acknowledgement

Transcripts used in this work were obtained under NCI R01-CA112379 (JEL) and NIMH R01-MH081098 (MTS). This work was also supported in part by the National Center for Research Resources and the National Center for Advancing Translational Sciences, National Institutes of Health (NIH), United States, through Grant UL1TR001414; and the National Institute of General Medical Sciences, NIH, through Grant T32-GM008620. The content is solely the responsibility of the authors and does not necessarily represent the official views of the NIH.

